# Durable SARS-CoV-2 B cell immunity after mild or severe disease

**DOI:** 10.1101/2020.10.28.20220996

**Authors:** Clinton O. Ogega, Nicole E. Skinner, Paul W. Blair, Han-Sol Park, Kirsten Littlefield, Abhinaya Ganesan, Pranay Ladiwala, Annukka AR Antar, Stuart C. Ray, Michael J. Betenbaugh, Andrew Pekosz, Sabra L. Klein, Yukari C. Manabe, Andrea L. Cox, Justin R. Bailey

**Affiliations:** Division of Infectious Diseases, Department of Medicine, Johns Hopkins University School of Medicine, Baltimore, Maryland, USA; W. Harry Feinstone Department of Molecular Microbiology and Immunology, The Johns Hopkins Bloomberg School of Public Health, Baltimore, Maryland, USA; Advanced Mammalian Biomanufacturing Innovation Center, Department of Chemical and Biomolecular Engineering, Johns Hopkins University, Baltimore, Maryland, USA

**Author notes:** **Address correspondence to:** Justin R. Bailey, MD, PhD, 855 N. Wolfe Street, Rangos Building, Suite 520, Baltimore MD 21205, USA. **Conflict of interest statement.** The authors declare that they have no conflicts of interest.

**Keywords:** SARS-CoV-2, COVID-19, neutralizing antibody, memory B cell

## Abstract

Multiple studies have shown loss of SARS-CoV-2 specific antibodies over time after infection, raising concern that humoral immunity against the virus is not durable. If immunity wanes quickly, millions of people may be at risk for reinfection after recovery from COVID-19. However, memory B cells (MBC) could provide durable humoral immunity even if serum neutralizing antibody titers decline. We performed multi-dimensional flow cytometric analysis of S protein receptor binding domain (S-RBD)-specific MBC in cohorts of ambulatory COVID-19 patients with mild disease, and hospitalized patients with moderate to severe disease, at a median of 54 (39-104) days after onset of symptoms. We detected S-RBD-specific class-switched MBC in 13 out of 14 participants, including 4 of the 5 participants with lowest plasma levels of anti-S-RBD IgG and neutralizing antibodies. Resting MBC (rMBC) made up the largest proportion of S-RBD-specific class-switched MBC in both cohorts. FCRL5, a marker of functional memory when expressed on rMBC, was dramatically upregulated on S-RBD-specific rMBC. These data indicate that most SARS-CoV-2-infected individuals develop S-RBD-specific, class-switched MBC that phenotypically resemble germinal center-derived B cells induced by effective vaccination against other pathogens, providing evidence for durable B cell-mediated immunity against SARS-CoV-2 after recovery from mild or severe COVID-19 disease.

**Graphical Abstract:** 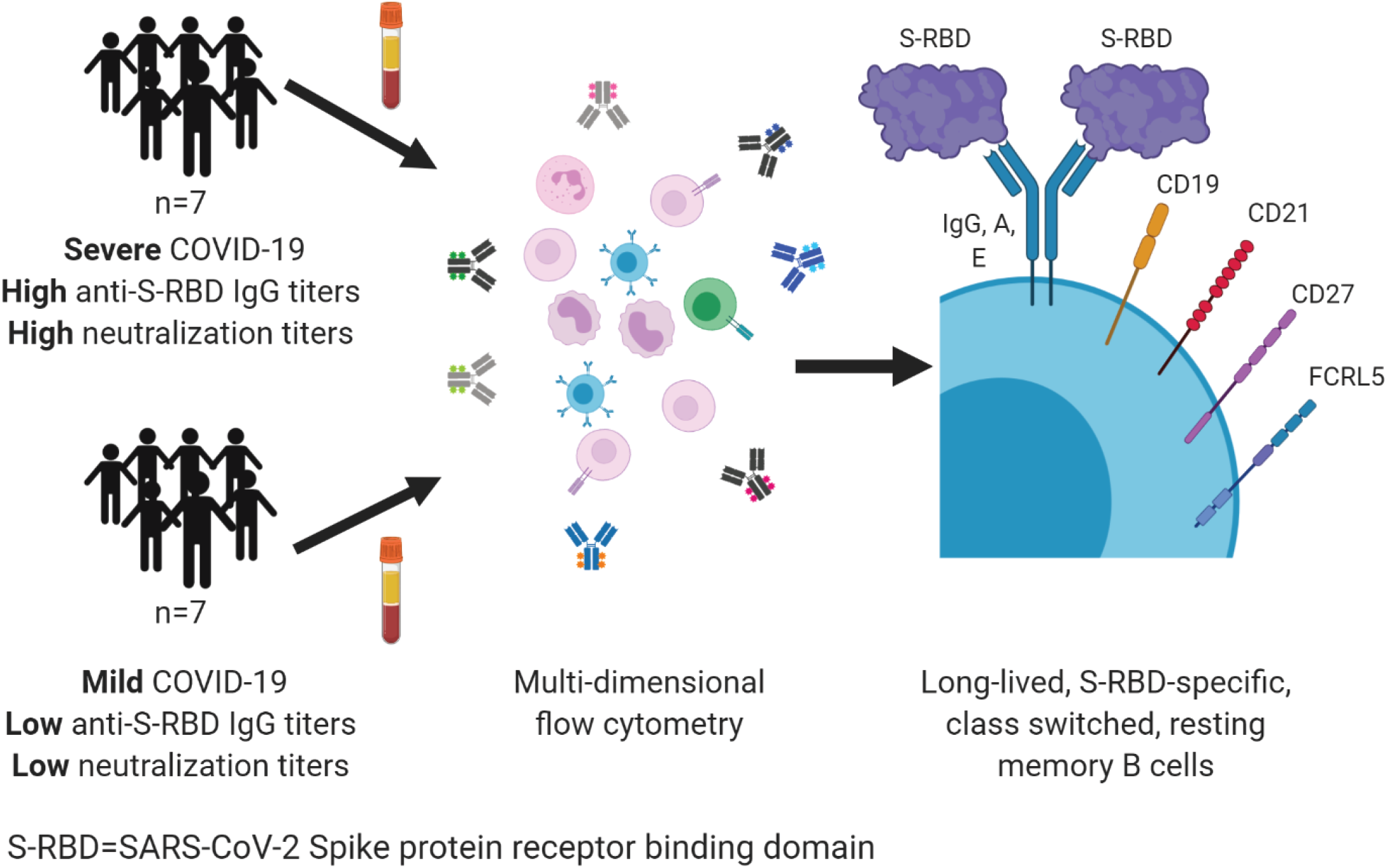

## Introduction

We are in the midst of an ongoing global pandemic caused by a novel coronavirus, SARS-CoV-2. COVID-19, the disease caused by SARS-CoV-2, can cause pulmonary inflammation, acute respiratory distress syndrome (ARDS), respiratory failure, and death. Despite the high morbidity and mortality caused by COVID-19, the majority of SARS-CoV-2-infected individuals recover and survive (1, 2). Following recovery, the durability of immunity against SARS-CoV-2 remains unclear. Durability of immunity is critical to mitigate the risk of reinfection for millions of people who have recovered or will recover from COVID-19.

After clearance of an infection or effective vaccination, phenotypically distinct B cell populations contribute to short- and long-term humoral immunity. Short-lived antibody-secreting cells (ASC) in blood and secondary lymphoid organs release antibodies into the circulation for weeks to months. Durable humoral immunity (lasting months to years) is mediated by bone marrow-resident, long-lived ASC and by memory B cells (MBC), which rapidly proliferate and differentiate into ASC in response to antigen re-challenge. Multiple studies have now demonstrated that serum antibody titers against SARS-CoV-2 wane and can even become undetectable after resolution of infection (3-6), likely reflecting a decline in short-lived ASC populations over time. Although other emerging reports have demonstrated more durable serum antibody responses (7-10), concerns remain that individuals who have recovered from COVID-19 may not maintain adequate immunity against reinfection. Individuals with mild COVID-19 disease generally mount lower titer antibody responses against the virus than those with severe disease (3, 10), raising particular concern that those who recover from mild infection are not protected against reinfection. If present and functional, MBC could provide durable humoral immunity even after the loss of detectable serum antibody titers, as has been demonstrated after vaccination against viruses like hepatitis B virus (11, 12). However, Kaneko et al. showed a dramatic loss of germinal centers during acute COVID-19, raising concern that T cell dependent, durable, class-switched SARS-CoV-2-specific MBC responses may not reliably develop after SARS-CoV-2 infection (13).

Little is known about the frequency and phenotype of SARS-CoV-2-specific MBC that develop in response to either severe or mild infection. B cells specific for the SARS-CoV-2 Spike (S) protein have been isolated from individuals with very low antibody titers, but the relatively low frequency of these cells has thus far limited further characterization (14). We developed a highly sensitive and specific flow cytometry-based assay to quantitate circulating SARS-CoV-2 S protein receptor binding domain (S-RBD)-specific B cells, and a cell surface phenotyping panel to characterize these cells. We focused on S-RBD-specific B cells because most virus-neutralizing human monoclonal antibodies target this domain (14-18). Neutralizing activity has been associated with protection against reinfection by other coronaviruses (19-22), and protection against challenge in animal models of SARS-CoV-2 infection (23, 24). Therefore, S-RBD-specific B cells are likely to be the cells responsible for production of protective neutralizing antibodies upon re-exposure.

Classical markers applied to these S-RBD-specific B cells allowed us to identify B cell lineages including non-class-switched B cells, class-switched ASC, class-switched resting (classical) MBC (rMBC), activated MBC (actMBC), atypical MBC (atyMBC), and intermediate MBC (intMBC). Additional subpopulations were identified by staining for a chemokine receptor (CXCR5), and potential inhibitory or activating receptors (FCRL5, CD22, and BTLA). Among the cell surface regulatory molecules, FCRL5 expression is of particular interest, since its expression on atyMBC has been associated with B cell dysfunction in chronic infections like HIV-1 and hepatitis C virus (25). In contrast, it is also upregulated on long-lived antigen-specific rMBC that develop after effective vaccination against influenza and tetanus (26, 27). This FCRL5+ rMBC population prefentially expands and forms plasmablasts on antigen re-challenge, indicating that FCRL5 expression on antigen-specific rMBC is a marker of effective long-lived B cell-mediated immunity.

To investigate the potential for durable B cell immunity after SARS-CoV-2 infection, we analyzed S-RBD-specific B cells in ambulatory COVID-19 patients with mild disease and hospitalized patients with moderate to severe disease. We detected S-RBD-specific non-class-switched B cells, S-RBD-specific class-switched ASC, and/or S-RBD-specific class-switched MBC in all participants, regardless of their serum antibody titers or disease severity. We analyzed the frequencies of these S-RBD-specific B cell populations, and of S-RBD-specific MBC subsets, including rMBC, intMBC, actMBC, and atyMBC. By also quantifying cell surface molecules CD38, FCRL5, CD22, BTLA, and CXCR5 on these MBC populations and subsets, we identified a phenotypic profile of S-RBD-specific class switched MBC that was consistent with functional, durable B cell immunity.

## Results

### Selection of study participants

B cells were obtained from participants with mild COVID-19 disease, moderate to severe disease, and from healthy COVID-19 negative controls (Table 1). Participants with mild COVID-19 disease who never required hospitalization or supplemental oxygen were identified in a previously described cohort of ambulatory patients (28). Symptoms in this cohort were tracked using a FLU-PRO score calculated from a participant survey, as previously described (28). To ensure that participants with mild disease were included in this study, a group of seven participants was selected with a median peak FLU-PRO score below the median peak score for the entire ambulatory cohort (FLU-PRO median (range)=0.09 (0.0-0.38) vs. 0.25 (0.0-1.63)). Seven additional participants with moderate to severe COVID-19 disease were selected from a second cohort of hospitalized patients (29), matched with the mild disease participants based on time since onset of symptoms at the time of blood sampling (median (range) time since symptom onset in days: ambulatory=61 (45-68); hospitalized=46 (39-104)). Peak supplemental oxygen support in hospitalized participants ranged from 2L via nasal cannula to mechanical ventilation. At the time of blood sampling for this study, five of the hospitalized participants had been discharged, and two remained hospitalized with critical illness. Hereafter, ambulatory, hospitalized, and healthy groups will be referred to as “mild”, “severe”, and “healthy”, respectively.

**Table 1.** Participant characteristics.

| Participant ID | Age<sup>1</sup> | Sex | Race | Ethnicity | Days from symptom onset | FLU-PRO<sup>2</sup> | Supp. O<sub>2</sub><sup>3</sup> |
| --- | --- | --- | --- | --- | --- | --- | --- |
| **Mild-ambulatory** |  |  |  |  |  |  |  |
| M1 | 60 | F | White | Non-Hispanic | 63 | 0.38 | none |
| M2 | 50 | F | Hawaiian/Pacific Islander | Non-Hispanic | 68 | 0.28 | none |
| M3 | 60 | F | White | Hispanic | 59 | 0.19 | none |
| M4 | 70 | F | White | Non-Hispanic | 64 | 0.06 | none |
| M5 | 60 | M | White | Non-Hispanic | 46 | 0 | none |
| M6 | 40 | M | Black | Non-Hispanic | 61 | 0.03 | none |
| M7 | 30 | M | American Indian/Alaska Native | Non-Hispanic | 45 | 0.09 | none |
| *median* | *60* |  |  |  | *61* | *0.09* |  |
| **Severe-hospitalized** |  |  |  |  |  |  |  |
| S1 | 70 | F | Black | Non-Hispanic | 90 | n/a | 4LNC |
| S2 | 50 | M | Black | Non-Hispanic | 104 | n/a | HFNC |
| S3 | 60 | M | White | Non-Hispanic | 48 | n/a | Intubated |
| S4 | 50 | M | Other | Non-Hispanic | 46 | n/a | 2LNC |
| S5 | 80 | F | Black | Non-Hispanic | 39 | n/a | Intubated |
| S6 | 50 | M | Black | Non-Hispanic | 42 | n/a | HFNC |
| S7 | 70 | M | Other | Non-Hispanic | 41 | n/a | Intubated |
| *median* | *60* |  |  |  | *46* |  |  |

<sup>1</sup>Age rounded to nearest 10 to avoid identifying participants. <sup>2</sup>Peak FLU-PRO score. <sup>3</sup>Maximum oxygen support required. 4LNC, 4 liters via nasal cannula; HFNC, high flow oxygen via nasal cannula; intubated, requiring mechanical ventilation; 2LNC, 2 liters via nasal cannula.

### Quantitation of S-RBD-specific B cells

A flow cytometry antibody panel was designed to identify non-class switched B cells (CD3-, CD19+, IgD/IgM+), class switched memory B cells (MBC) (CD3-, CD19+, IgM-, IgD-, CD38+/− (excluding ++), CD138-) and class switched antibody secreting cells (ASC) (CD3-, CD27+, CD19+/−, IgM-, IgD-, CD38++) (Supp. Figure 1). The frequency of all non-class switched B cells, class switched MBC, or class switched ASC among single viable lymphocytes was not significantly different between healthy, mild, and severe groups, but there was a trend toward greater frequency of class-switched ASC in the severe group compared to mild and healthy groups (Figure 1A). As we defined these three B cell populations, we used a 6x-histidine tagged, soluble S-RBD protein followed by anti-His Alexa Fluor 647-conjugated antibody to stain cells expressing S-RBD-specific antibodies on their surface (Figure 1B and Supplemental Figure 1). We quantitated the frequency of these S-RBD-specific cells among non-class switched B cells, class switched ASC, and class switched MBC (Figure 1C). Four of seven (57%) mild and seven of seven (100%) severe participants had a frequency of S-RBD-specific non-class switched B cells above the true positive threshold set using the healthy group. The frequency of these cells did not differ significantly between the mild and severe groups. Four of seven (57%) mild and four of seven (57%) severe participants had a frequency of S-RBD-specific class switched ASC above the true positive threshold. The frequency of these cells also did not differ significantly between the mild and severe groups. Six of seven (86%) mild and seven of seven (100%) severe participants had a frequency of S-RBD-specific class switched MBC above the true positive threshold. The single individual without detectable S-RBD-specific class switched MBC was asymptomatic throughout infection (peak FLU-PRO=0.0). Frequency of S-RBD-specific class switched MBC was significantly higher in severe participants than in mild participants (mean S-RBD+ frequency 0.85% vs. 0.20%, p=0.004). Taken together, these data demonstrate that S-RBD-specific cells could be detected among non-class switched B cells and class switched ASC in most SARS-CoV-2-infected participants, and S-RBD-specific class switched MBC could be detected in thirteen of fourteen participants. S-RBD-specific cells were significantly more frequent among class switched MBC from the severe group relative to the mild group.

**Figure 1.**
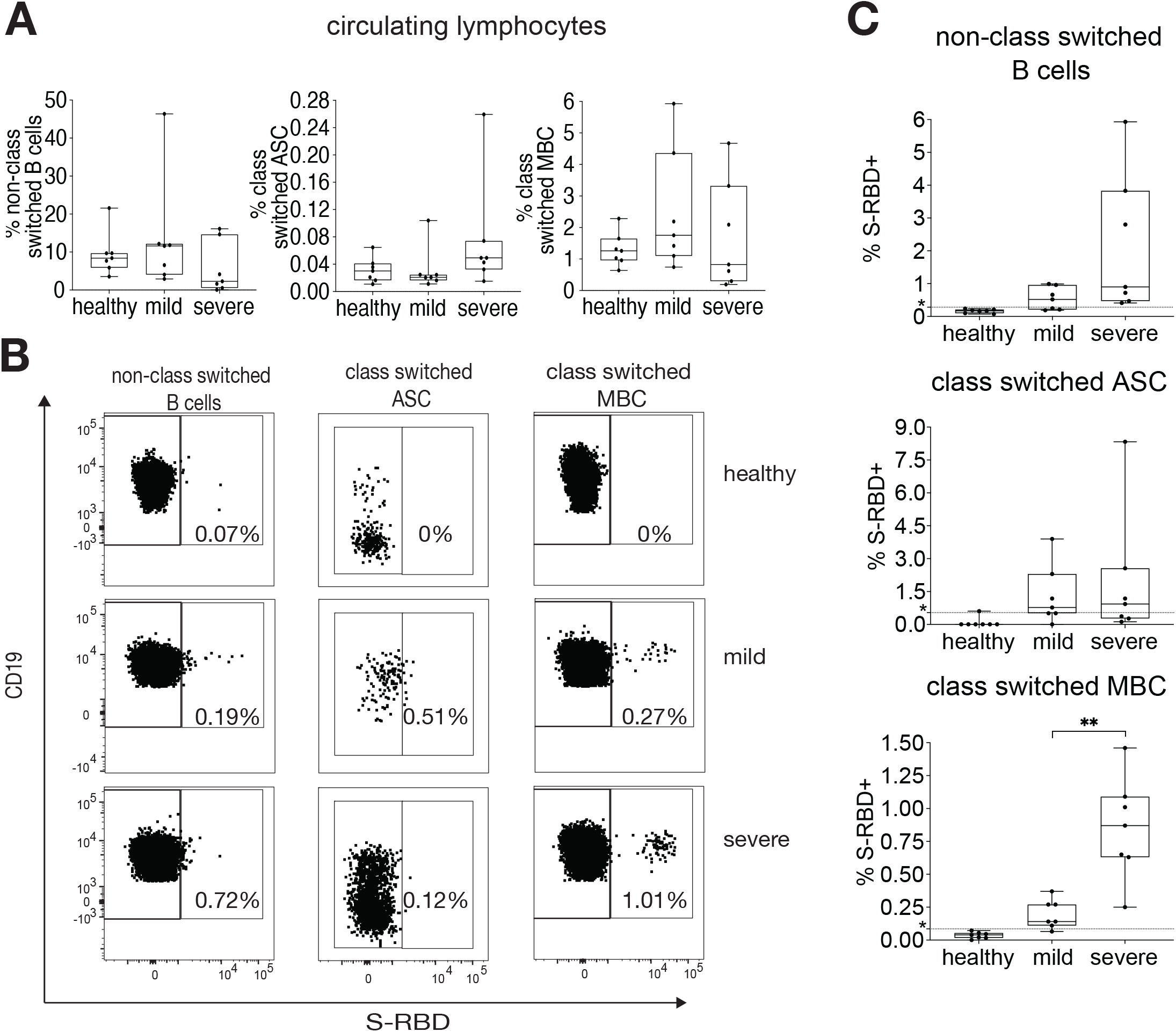
Quantifying S-RBD specific B cells. **(A)** % of lymphocytes that are class switched MBC, class switched ASC, or non-class switched B cells in healthy, mild, and severe participants (N=7 for each group). **(B)** Gating strategy for S-RBD specific non-class switched B cells (CD3-, CD19+, IgD/IgM+, S-RBD+), S-RBD specific class switched MBC (CD3-, CD19+, IgM-, IgD-, CD38+/− (excluding ++), CD138-, S-RBD+), and S-RBD specific class switched ASC (CD3-, CD19+/−, IgM-, IgD-, CD38++, CD27+, S-RBD+) in “healthy” (COVID-19 negative), “mild” (COVID-19+, ambulatory), and “severe” (COVID-19+, hospitalized) participants. **(C)** % of class switched MBC, class switched ASC, and non-class switched B cells that are S-RBD specific in healthy, mild, and severe participants (N=7 for each group). Dotted line represents the true positive threshold, defined as the mean plus two standard deviations of the healthy group. For **A and C** box plots, horizontal lines indicate means, boxes are inter-quartile range, and whiskers are minimum to maximum. Three group comparisons in **A** were performed using one-way ANOVA, with p values adjusted for multiple comparisons using the Benjamini, Krieger and Yekutieli method. Two group comparisons in **C** were performed with t tests if data were normally distributed based on Shapiro Wilk normality test or Mann Whitney test if data were not normally distributed. Statistically significant comparisons are indicated (** P ≤ 0.01)

### Detectable S-RBD-specific MBC despite low levels of anti-S-RBD IgG and neutralizing antibodies in plasma

Given concerns that low or waning plasma titers of neutralizing antibodies in some individuals indicate a lack of a durable humoral response, we were interested in evaluating whether COVID-19 participants with low levels of plasma anti-S-RBD IgG and low neutralizing antibody levels had detectable S-RBD-specific MBC in circulation. S-RBD binding IgG was measured using serial dilutions of plasma in an ELISA, and neutralizing antibodies were measured with serial dilutions of plasma in a microneutralization assay using replication competent SARS-CoV-2 virus (10). Curves were fit to these data, and area under the curve (AUC) values calculated. Anti-S-RBD IgG and neutralization AUC values each varied over a wide range across study subjects (1e2.7-1e4.9 and 1e0.8-1e3.0, respectively). As expected based on prior studies (3, 10), there was a trend toward higher anti-S-RBD IgG and neutralization AUC values in the severe group relative to the mild group, although these differences were not statistically significant, likely due to the small number of subjects (Figure 2A-B). We next evaluated whether we could detect S-RBD-specific cells among class switched MBC of participants with low, intermediate, or high levels of plasma anti-S-RBD IgG (Figure 2C) or low, intermediate, or high levels of neutralizing antibodies (Figure 2D). We detected S-RBD-specific class switched MBC above the nonspecific background frequency in 4 of 5 participants with the lowest IgG and neutralization AUCs, 5 of 5 with intermediate AUCs, and 4 of 4 with highest AUCs. The single individual without detectable S-RBD-specific class switched MBC had the lowest levels of plasma anti-S-RBD IgG (AUC=1e2.7) and neutralizing antibodies (AUC=1e0.8) in the study. There was a non-significant trend toward higher frequencies of RBD specific cells among class switched MBC in the higher AUC IgG and higher neutralizing antibody AUC groups relative to the lowest AUC groups. Overall, these data show that S-RBD-specific class switched MBC were detectable in the circulation of most infected individuals, including the majority of subjects with low levels of plasma anti-S-RBD IgG and neutralizing antibodies.

**Figure 2.**
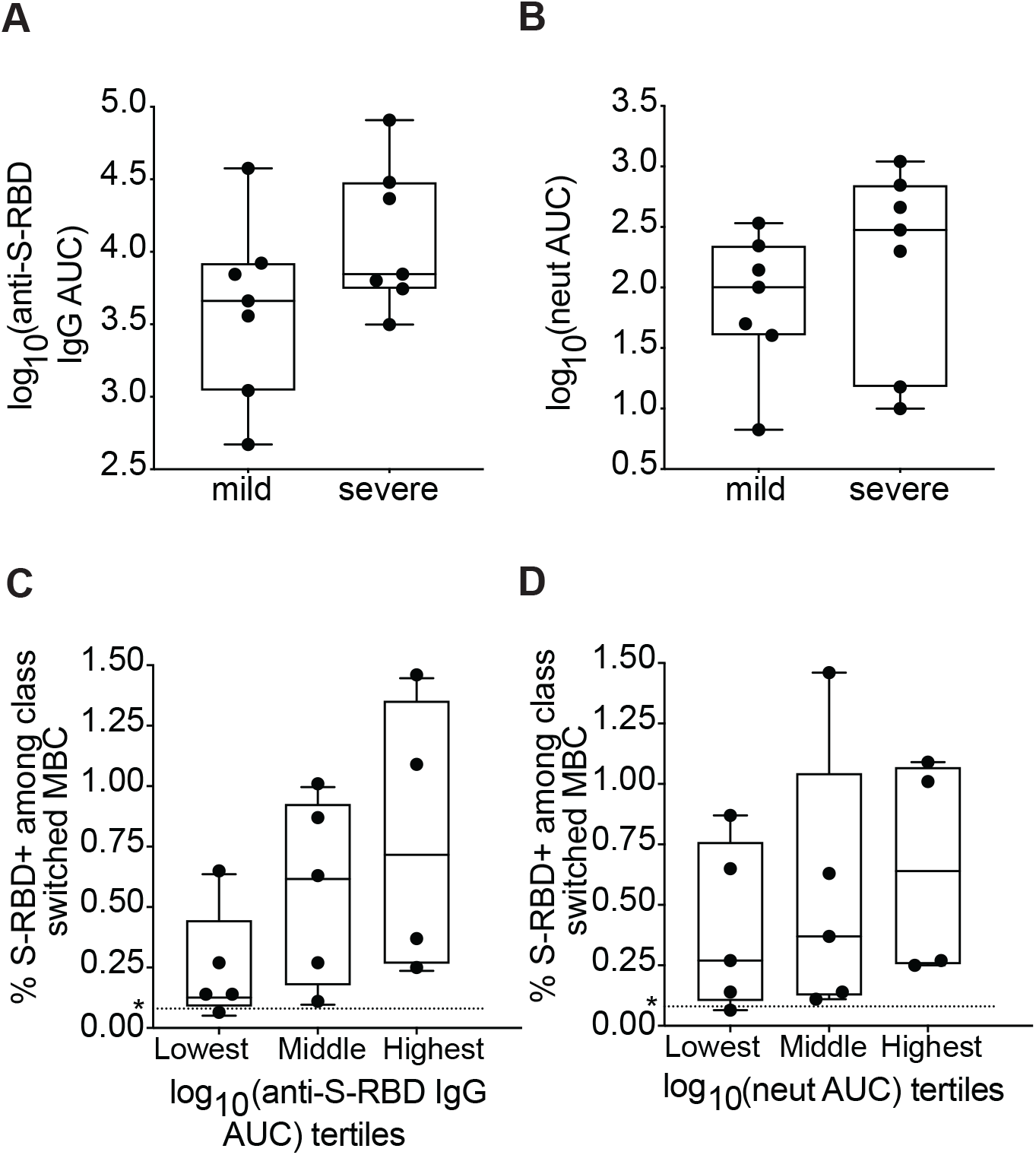
Comparisons of serum anti S-RBD IgG and neutralizing antibody titers in mild and severe participants. **(A)** Anti S-RBD IgG area under the curve (AUC) in mild and severe participants. **(B)** Neutralizing antibody AUC in mild and severe participants. **(C)** % of class switched MBC that are RBD specific, from participants with plasma anti-S-RBD AUC values in the lowest, middle, and highest tertiles. **(D)** % of class switched MBC that are RBD specific, from participants with plasma neutralizing antibody AUC values in the lowest, middle, and highest tertiles. Dotted line represents the true positive threshold, defined as the mean plus two standard deviations of the healthy group. For box plots, horizontal lines indicate means, boxes are inter-quartile range, and whiskers are minimum to maximum. P values for **A-B** were calculated using t tests and P values for **C-D** were calculated using one-way ANOVA test for linear trend. No statistically significant comparison was found.

### UMAP analysis of class switched MBC surface markers

To further characterize the phenotypes of S-RBD-specific and nonspecific class switched MBC in healthy, mild, or severe COVID-19 patients, we studied surface expression of CD21, CD27, FCRL5, CXCR5, CD22, BTLA, and CD38. CD21 and CD27 expression allow separation of MBC into intermediate MBC (intMBC, CD21+ CD27-), resting MBC (rMBC, CD21+, CD27+), activated MBC (actMBC, CD21-CD27+), and atypical MBC (atyMBC, CD21-CD27-) subsets. B- and T-lymphocyte attenuator (BTLA) or CD272 and CD22/Siglec2 are immune cell inhibitory receptors with cytoplasmic immunoreceptor tyrosine-based inhibition motifs (ITIMs) (30-33), while FCRL5 has two ITIMs and one immunoreceptor tyrosine-based activation motif (ITAM) (34, 35). C-C chemokine receptor type 5 (CXCR5) is a germinal center homing receptor that is generally down-regulated after cells have undergone class switching and somatic hypermutation (36, 37). CD38 has relatively undefined functional significance, but it is negative on “late” class-switched MBC and positive on “early” class switched MBC that emerged more recently from germinal center reactions (38).

We first analyzed a UMAP projection of all class switched MBC from healthy, mild, and severe groups generated based on binding of S-RBD and expression of CD21, CD27, CD38, CD22, FCRL5, CXCR5, and BTLA (Figure 3A). This UMAP identified a clear segregation of co-mingled mild and severe S-RBD+ cells from RBD- and healthy donor cells. From this UMAP clustering projection, we extrapolated multigraph color mapping of the receptors with observed differential expression of all surface markers except BTLA (Figure 3B). To determine whether there was any shift in expression of these markers between study groups, we overlayed expression histograms of all the surface receptors of healthy, mild S-RBD-, mild S-RBD+, severe S-RBD-, and severe S-RBD+ class switched MBC (Figure 3C). Again, there was no observable shift in the expression of BTLA. For all study groups, there was a bimodal distribution of CD21 expression, indicating that both CD21+ (intMBC and rMBC) and CD21-(actMBC and atyMBC) MBC subsets were present. For both mild and severe S-RBD+ MBC, we observed a shift toward increased expression of CD22 and FCRL5 and downregulation of CXCR5 compared to healthy and S-RBD-populations. For CD38, we observed upregulation in mild and severe S-RBD+ MBC and downregulation in mild and severe S-RBD-MBC compared to healthy participants.

**Figure 3:**
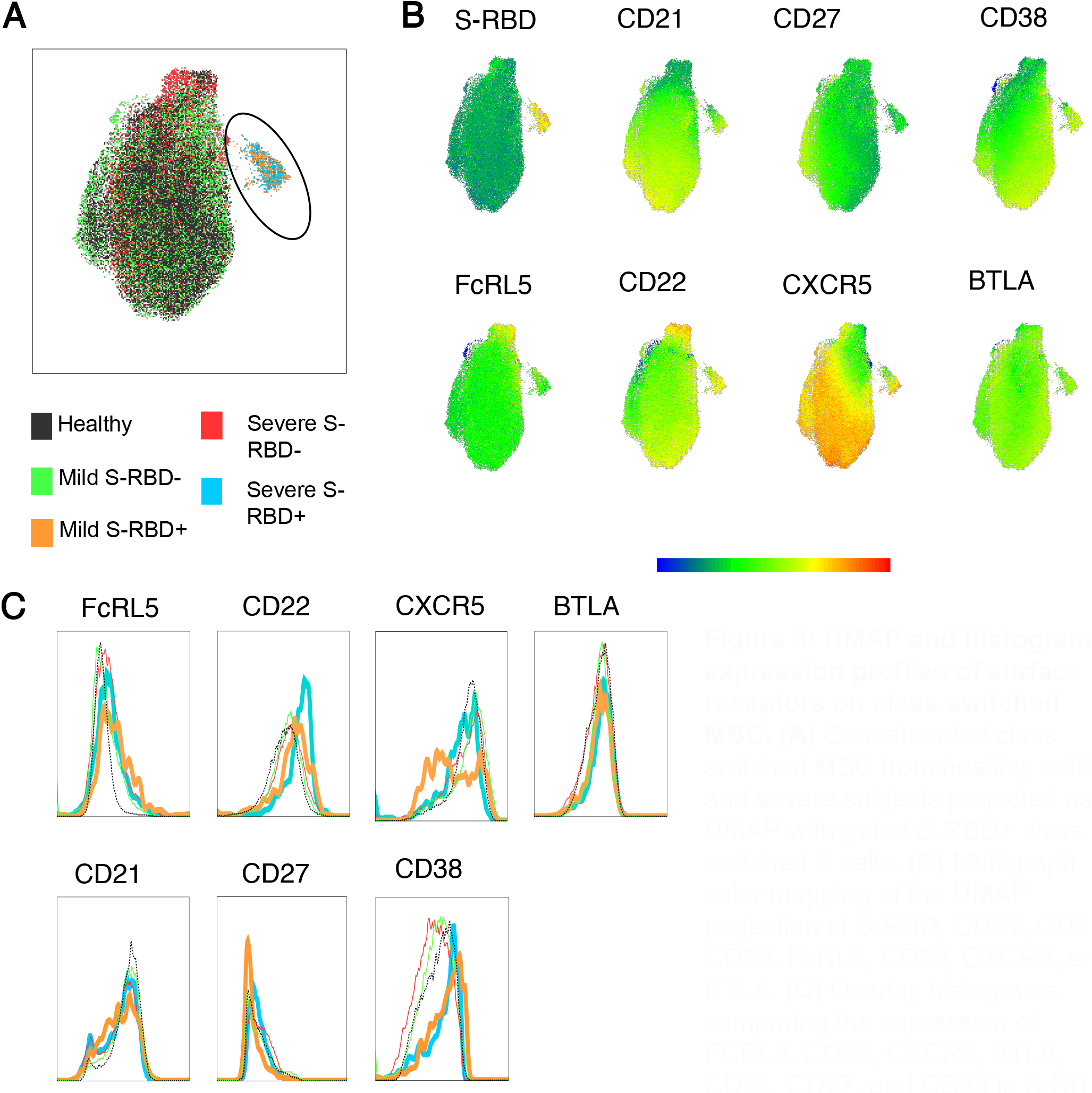
UMAP and histogram expression profiles of surface receptors on class switched MBC. **(A)** Concatenated class switched MBC from healthy, mild, and severe subjects projected as UMAP with gated S-RBD+ class switched B cells. **(B)** Multigraph color mapping of the UMAP projection of S-RBD, CD21, CD27, CD38, FcRL5, CD22, CXCR5, and BTLA. **(C)** Overlay histograms comparing the expression of FCRL5, CD22, CXCR5, BTLA, CD21, CD27, and CD38 in S-RBD- or S-RBD+ healthy, mild and severe groups.

### Quantifying subsets of S-RBD nonspecific and S-RBD-specific class switched MBC

To better understand the functional phenotypes of the S-RBD-specific MBC identified in both mild and severe groups, we compared the frequencies of intMBC, rMBC, actMBC, and atyMBC among S-RBD-specific and S-RBD nonspecific class switched MBC at the level of individual participants (Figure 4). MBC subsets identified based on CD21 and CD27 expression have notably different phenotypes (38). Classical MBC, also called rMBC, persist for months to years and respond to antigen re-challenge by proliferating and differentiating into antibody-producing ASC. ActMBC are cells that recently left germinal centers and are already primed to become antibody secreting plasma cells (39). AtyMBC are generally thought to be poorly functional, and are present at higher frequencies in chronic infections like HIV-1, hepatitis C virus, tuberculosis, or malaria (25, 40, 41). AtyMBC were also found to be more frequent among bulk (not antigen specific) MBC during acute SARS-CoV-2 infection (42). IntMBC likely represent a transitional state between MBC subsets.

**Figure 4:**
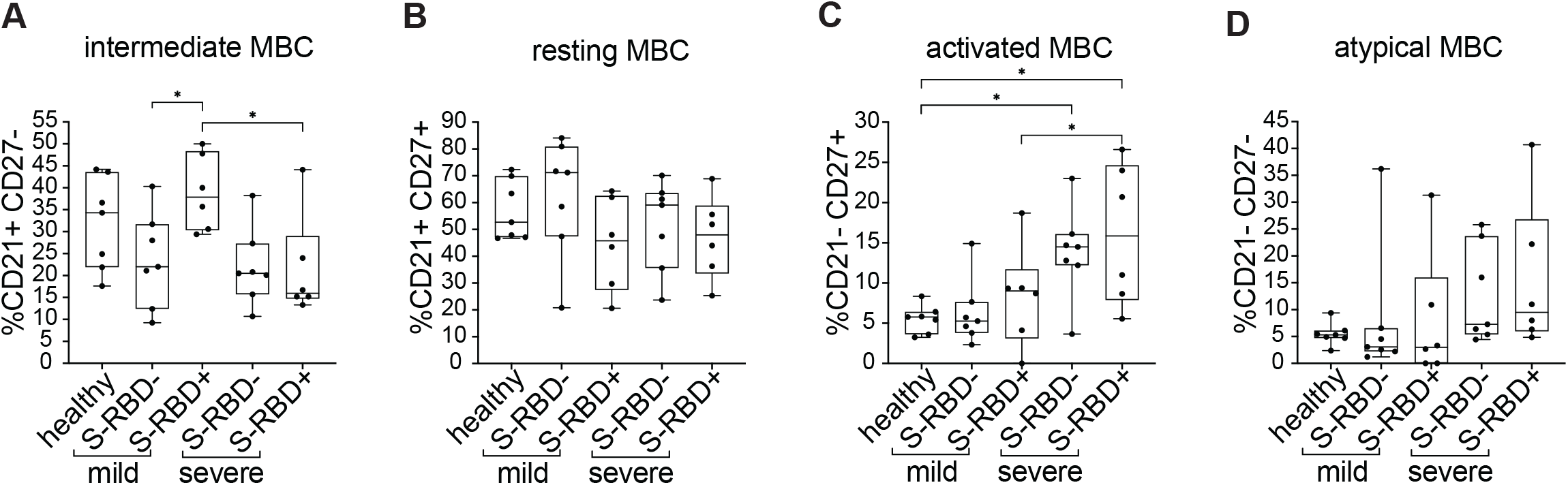
Frequency of MBC subsets in S-RBD nonspecific (S-RBD-) or S-RBD specific (S-RB-D+) class switched MBC from healthy, mild, or severe participants. **(A)** intMBC (CD21+, CD27-). **(B)** rMBC (CD21+, CD27+). **(C)** actMBC (CD21-, CD27+). **(D)** atyMBC (CD21-, CD27-). For box plots, horizontal lines indicate means, boxes are inter-quartile range, and whiskers are minimum to maximum. Groups were compared by one-way ANOVA with p values adjusted for multiple comparisons using the Benjamini, Krieger and Yekutieli method. Statistically significant comparisons are indicated (* P ≤ 0.05).

S-RBD-specific class switched MBC from two participants were not included in these or subsequent analyses, either because they did not have detectable S-RBD-specific class switched MBC frequency above background (subject M5), or because absolute cell numbers were low given severe lymphopenia (subject S3). S-RBD nonspecific MBC were adequately abundant in all participants to allow their inclusion in all analyses (see Methods). There were no statistically significant differences in the frequencies of intMBC, rMBC, or atyMBC subsets among S-RBD-specific or S-RBD nonspecific class switched MBC from healthy, mild or severe participants (Figure 4). In contrast, actMBC were significantly more frequent among both S-RBD nonspecific and S-RBD-specific MBC populations in severe participants compared to healthy and mild participants (e.g. mean frequency of severe S-RBD+ MBC vs. healthy S-RBD-MBC, 16.09% vs. 5.53%, p=0.01) (Figure 4C). This likely represents greater ongoing immune activation in the severe infection group relative to the healthy and mild groups and is also consistent with the observed trend toward higher frequency of ASC in the severe group (Figure 1A). Overall, these data demonstrate an expected distribution of S-RBD-specific cells among MBC subsets, with the largest proportion of S-RBD-specific class switched MBC in both mild and severe groups falling in the rMBC subset.

### Expression of activating or inhibitory surface markers on class switched MBC and MBC subsets

To further investigate the differential expression of surface markers that we observed in the UMAP projections and histograms of grouped samples, we compared expression of FCRL5, CXCR5, CD22, and CD38 at the level of individual participants between healthy, mild S-RBD-, mild S-RBD+, severe S-RBD-, and severe S-RBD+ groups (Figure 5). BTLA expression was not included in this analysis given no differential expression in the UMAP. We found that FCRL5 was dramatically upregulated in mild S-RBD+ MBC relative to healthy cells, mild S-RBD-cells, and severe S-RBD+ cells (p=<0.0001, 0.003, and 0.01, respectively). FCRL5 was also upregulated to a lesser, but still significant extent on severe S-RBD+, mild S-RBD-, and severe S-RBD-MBC relative to healthy MBC (p=0.01, 0.03, and 0.04, respectively) (Figure 5A). The frequency of CXCR5+ cells among class switched MBC was not significantly different between the groups, although it was downregulated in 3 of 6 participants in both mild and severe groups (Figure 5B) which could account for the shift observed in the histogram in Figure 3C. Since CD22/siglec-2 is ubiquitously expressed on B cells, we analyzed its relative expression by comparing mean fluorescence intensities (MFI). Compared to healthy controls, CD22 was upregulated on mild S-RBD+ class switched MBC (p=0.04) (Figure 5C). Among class switched MBC, CD38 expression did not differ significantly between SARS-CoV-2 infected and healthy participants (Figure 5D).

**Figure 5:**
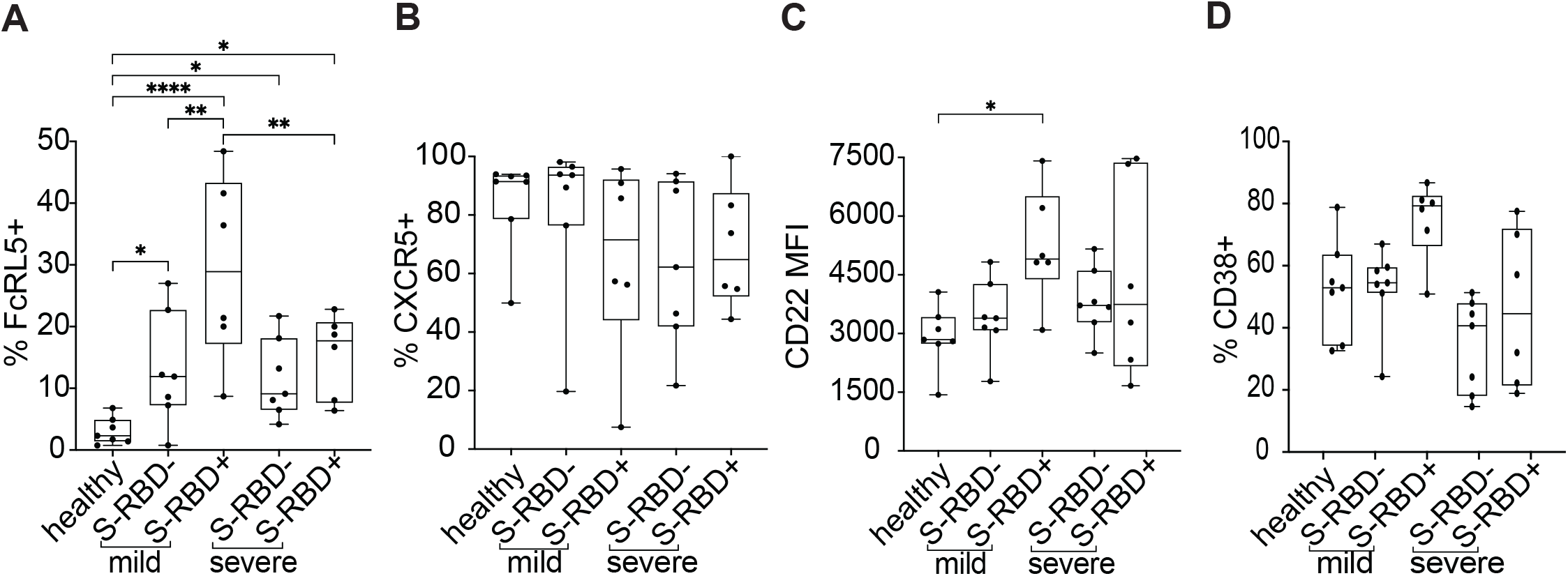
Surface expression of FcRL5, CXCR5, CD22, and CD38 on S-RBD nonspecific (S-RBD-) or S-RBD specific (S-RBD+) class switched MBC from healthy, mild, or severe participants. Expression is shown as either percent of cells positive or the mean fluorescent intensity (MFI). **(A)** FcRL5 **(B)** CXCR5 **(C)** CD22 **(D)** CD38. For box plots, horizontal lines indicate means, boxes are inter-quartile range, and whiskers are minimum to maximum. Groups were compared by one-way ANOVA with p values adjusted for multiple comparisons using the Benjamini, Krieger and Yekutieli method. Statistically significant comparisons are indicated (* P ≤ 0.05, ** P ≤ 0.01, *** P ≤ 0.001, **** P ≤ 0.0001).

Having observed upregulation of both FCRL5 and CD22, a trend toward downregulation of CXCR5, and a trend toward upregulation of CD38 on S-RBD+ class switched MBC, we analyzed expression of these surface markers on MBC subsets rMBC, intMBC, actMBC, and atyMBC (Figure 6). As with total class switched RBD+ MBC, we found that FCRL5 was dramatically upregulated on mild S-RBD+ rMBC relative to healthy rMBC, mild S-RBD-rMBC, and severe S-RBD+ rMBC (p=<0.0001, <0.0001, and <0.0001, respectively). FCRL5 was also upregulated to a lesser, but still significant extent on severe S-RBD+ and mild S-RBD-rMBC relative to healthy rMBC (p=0.02 and 0.01, respectively). CXCR5 was significantly downregulated on mild S-RBD+ atyMBC relative to healthy atyMBC, mild S-RBD-atyMBC, and severe S-RBD+ atyMBC (p=0.001, 0.003, and 0.001, respectively). CD22 was significantly upregulated on mild S-RBD+ rMBC and intMBC relative to healthy cells (p=0.02 and 0.03, respectively). CD38+ cells were significantly more frequent among mild S-RBD+ actMBC relative to healthy actMBC and mild S-RBD-actMBC (p=0.007 and 0.01, respectively). Taken together, these results indicate that FCRL5 was significantly upregulated on S-RBD-specific rMBC in both mild and severe infection. In mild but not severe infection, CD22 was upregulated on S-RBD-specific rMBC and intMBC, CXCR5 was downregulated on S-RBD-specific atyMBC, and CD38 was upregulated on S-RBD-specific actMBC.

**Figure 6:**
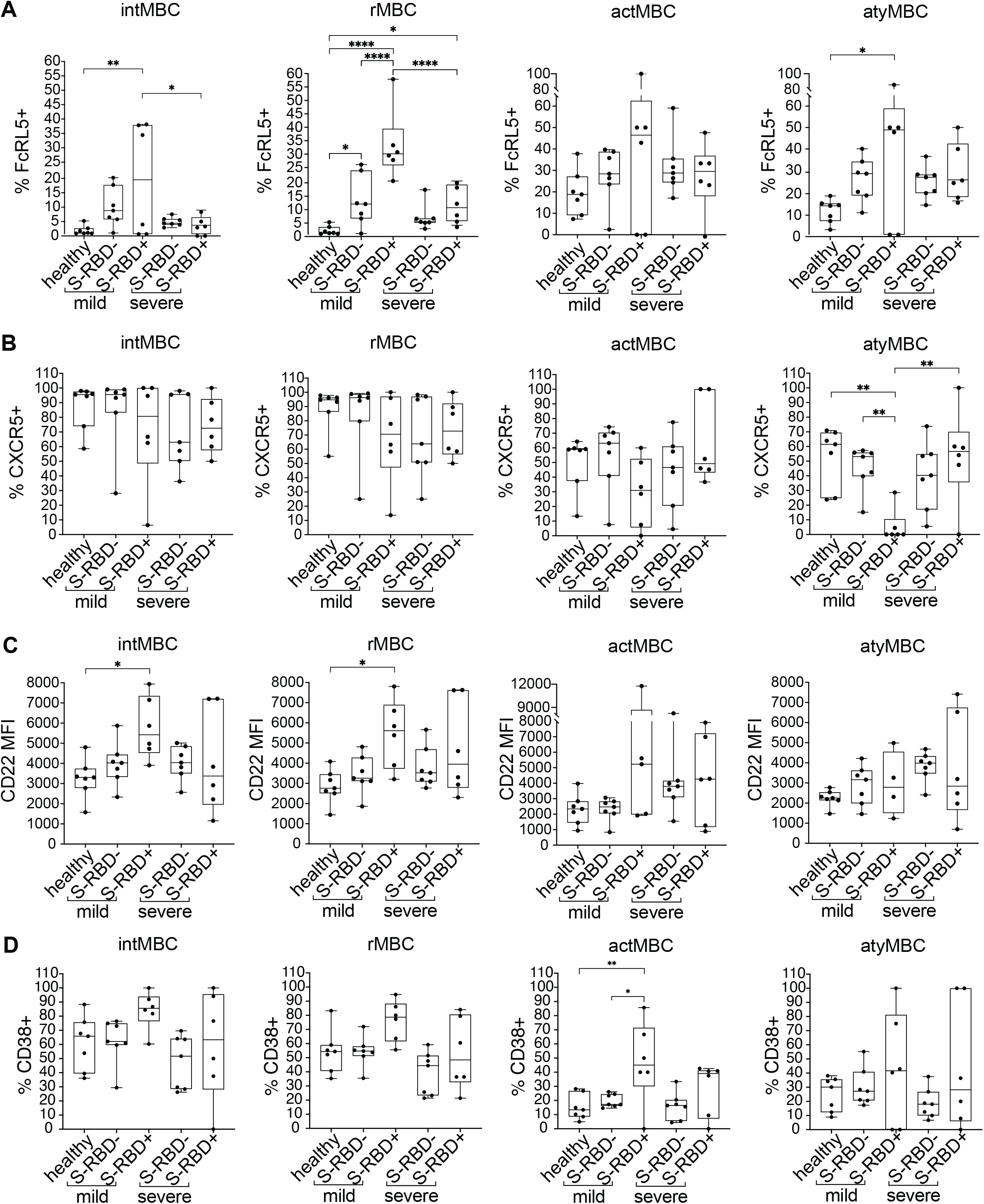
Surface expression of FcRL5, CXCR5, CD22, and CD38 on S-RBD nonspecific (S-RBD-) or S-RBD specific (S-RBD+) class switched intMBC, rMBC, actMBC, or atyMBC from healthy, mild, or severe participants. Expression is shown as either percent of cells positive or the mean fluorescent intensity (MFI). **(A)** FcRL5 **(B)** CXCR5 **(C)** CD22 **(D)** CD38. For box plots, horizontal lines indicate means, boxes are inter-quartile range, and whiskers are minimum to maximum. Groups were compared by one-way ANOVA with p values adjusted for multiple comparisons using the Benjamini, Krieger and Yekutieli method. Statistically significant comparisons are indicated (* P ≤ 0.05, ** P ≤ 0.01, *** P ≤ 0.001, **** P ≤ 0.0001).

## Discussion

To investigate the durability of B cell immunity after SARS-CoV-2 infection, we analyzed S-RBD-specific B cells in ambulatory COVID-19 patients with mild disease and hospitalized patients with moderate to severe disease, at a median of 54 days after onset of symptoms. We detected S-RBD-specific class-switched MBC in 13 out of 14 participants, including 4 of the 5 participants with lowest plasma levels of anti-S-RBD IgG and neutralizing antibodies. The largest proportion of S-RBD-specific class-switched MBC in both cohorts were rMBC. AtyMBC, which tend to be poorly functional, were a minor population. FCRL5 was upregulated on S-RBD-specific rMBC after severe infection, and upregulated even more dramatically after mild infection.

These findings are of particular interest given the observation of Kaneko et al. of a dramatic loss of germinal centers in lymph nodes and spleens after SARS-CoV-2 infection (13). This observation would suggest that SARS-CoV-2-specific B cells in infected individuals lack T cell help and would therefore have reduced capacity to undergo class switching and transition to a resting memory phenotype. Our data indicate that despite this loss of germinal centers, T cell help is adequate to facilitate class switching of S-RBD-specific B cells, and transition of many of these cells to a resting state, regardless of disease severity. We did not measure the extent of somatic hypermutation of these B cells, but multiple groups have already demonstrated that human S-RBD-specific antibodies acquire enough somatic mutations to achieve very high affinity (14-18), again demonstrating that T cell help is adequate in most individuals.

A prior study by Oliviero et al. of bulk (not antigen-specific) MBC subsets during acute or convalescent COVID-19 found that atyMBC were expanded during acute infection, with atyMBC frequencies normalizing during convalescence (42). Our study extends that evaluation by studying both S-RBD-specific and S-RBD nonspecific MBC. We found that S-RBD-specific and S-RBD nonspecific atyMBC frequencies did not differ significantly from healthy controls, but S-RBD-specific actMBC were expanded in severely infected individuals. The contrast of our results with those of Oliviero et al. likely arise from differing timing after infection, and also by our focus on antigen-specific MBC. The fact that atyMBC make up a small minority of S-RBD-specific MBC provides further evidence that most S-RBD-specific MBC are normally functional.

It is interesting that the single individual without detectable S-RBD-specific class switched MBC was asymptomatic throughout infection, and also had the lowest levels of anti-S-RBD IgG and neutralizing antibodies in the study. It is possible that those with asymptomatic disease, who tend to develop lower neutralizing antibody titers, may also develop lower frequencies of class switched RBD-specific MBC. Given the low frequency of RBD specific MBC across the cohort, we would need to analyze a larger number of PBMC to confirm with confidence that this individual is truly negative for S-RBD-specific class switched MBC.

A limitation of this study is the lack of long-term longitudinal sampling of B cells after infection, which would be required to prove that the S-RBD-specific MBC responses observed here are truly durable. These studies will be pursued as longitudinal samples become available. However, we have shown here that S-RBD-specific MBC in most infected individuals have a phenotype that very closely resembles the phenotype of germinal center-derived MBC induced by effective vaccination against influenza and tetanus. Of particular note is the upregulation of FCRL5 on S-RBD-specific class switched rMBC after either mild or severe disease. FCRL5 is expressed by most germinal center-derived MBC in plasmodium-infected mice, and these FCRL5+ MBC differentiate into ASC on re-challenge (26). In addition, Kim et al. found that in humans, presumably vaccinated against tetanus months to years prior, FCRL5 was upregulated on tetanus specific rMBC (CD21+, CD27+) but not on bulk rMBC (26). Nellore et al. showed similar results after influenza vaccination of humans, demonstrating that hemagglutinin (HA)-specific, FCRL5+ MBC were induced by vaccination, and that these FCRL5+ MBC preferentially differentiated into plasmablasts upon antigen rechallenge approximately a year after vaccination (27). Further studies will be necessary to understand the implications of CD22 upregulation on S-RBD-specific rMBC and intMBC, CXCR5 downregulation on S-RBD-specific atyMBC, and CD38 upregulation on S-RBD-specific actMBC in the mild group but not the severe group, but none of these differences necessarily indicate dysfunction in either group. Overall, despite our lack of longitudinal testing, the phenotypic similarity of S-RBD-specific MBC in this study to typical, germinal center-derived MBC induced by effective vaccination provide strong evidence that these S-RBD-specific MBC are durable and functional.

In summary, we have demonstrated that S-RBD-specific class-switched MBC develop in most SARS-CoV-2-infected individuals, including those with mild disease or low levels of plasma anti-S-RBD IgG and neutralizing antibodies. The most abundant subset of S-RBD-specific class-switched MBC in both cohorts were rMBC, and atyMBC were a minor population. FCRL5, a marker of a functional memory response when expressed on antigen-specific rMBC, was dramatically upregulated on S-RBD-specific rMBC, particularly after mild infection. These data indicate that most SARS-CoV-2-infected individuals develop S-RBD-specific, class-switched MBC that phenotypically resemble B cells induced by effective vaccination against other pathogens, providing evidence for durable humoral immunity against SARS-CoV-2 after recovery from either mild or severe COVID-19 disease. These data have implications for risk of reinfection after recovery from COVID-19, and also provide a standard against which B cell responses to novel SARS-CoV-2 vaccines could be compared.

## Methods

### Study participants

Participants with mild COVID-19 disease who never required hospitalization or supplemental oxygen were identified in a cohort of ambulatory COVID-19 patients. Symptoms in this cohort were tracked using a FLU-PRO score calculated from a participant survey, as previously described (28). Participants with moderate to severe COVID-19 disease were selected from a cohort of hospitalized patients (29), matched with the mild participants based on time since onset of symptoms at the time of blood sampling. PBMC cryopreserved prior to the onset of the COVID-19 pandemic were also obtained from anonymous healthy blood donors. Healthy and COVID-19 participant blood specimens were ficoll gradient separated into plasma and PBMCs. PBMCs were viably cryopreserved in FBS + 10% DMSO for future use.

### Expression and purification of soluble Spike protein Receptor Binding Domain (S-RBD)

#### Plasmid preparation

Recombinant plasmid constructs containing modified S protein Receptor Binding Domain (S-RBD) and a beta-lactamase (amp) gene were obtained (Stadlbauer 2020) and amplified in E.coli after transformation and growth on LB agar plates coated with Ampicillin. The plasmids were extracted using GigaPrep kits (Thermo Fisher Scientific) and eluted in molecular biology grade water.

#### Recombinant protein expression

HEK293.2sus cells (ATCC) were obtained and adapted to Freestyle™ F-17 medium (Thermo Fisher Scientific) and BalanCD® (Irvine Scientific) using polycarbonate shake flasks (Fisherbrand) with 4mM GlutaMAX supplementation (Thermo Fisher Scientific). The cells were routinely maintained every 4 days at a seeding density of 0.5 million cells/mL. They were cultured at 37°C, 90% humidity with 5% CO2 for cells in BalanCD® while those in F-17 were maintained at 8% CO2. Cells were counted using trypan blue dye (Gibco) exclusion method and a hemocytometer. Cell viability was always maintained above 90%. Twenty-four hours prior to transfection (Day -1), the cells were seeded at a density of 1 million cells/mL, ensuring that the cell viability was above 90%. Polyethylenimine (PEI) stocks, with 25 kDa molecular mass (Polysciences), were prepared in MilliQ water at a concentration of 1 mg/mL. This was filter sterilized through a 0.22 μm syringe filter (Corning), aliquoted and stored at −20°C. On the day of transfection (Day 0), the cells were counted to ensure sufficient growth and viability. OptiPRO™ SFM (Gibco) was used as the medium for transfection mixture. For 100 mL of cell culture, 2 tubes were aliquoted with 6.7 mL each of OptiPRO™, one for PEI and the other for rDNA. DNA:PEI ratio of 1:3.5 was used for transfection. A volume of 350 μL of prepared PEI stock solution was added to tube 1 while 100 μg of rDNA was added to tube 2 and incubated for 5 minutes. Post incubation, these were mixed together, incubated for 10 minutes at RT and then added to the culture through gravity addition. The cells were returned back to the 37°C incubator. A day after transfection (Day 1), the cells were spun down at 1,000 rpm for 7 minutes at RT and resuspended in fresh media with GlutaMAX™ supplementation. 3-5 hours after resuspension, 0.22 μm sterile filtered Sodium butyrate (EMD Millipore) was added to the flask at a final concentration of 5 mM (Grünberg et al.). The cells were allowed to grow for a period of 4-5 days. Cell counts, viability, glucose and lactate values were measured every day. Cells were harvested when either the viability fell below 60% or when the glucose was depleted, by centrifugation at 5000 rpm for 10 minutes at RT. Cell culture supernatants containing either recombinant S-RBD or S protein were filtered through 0.22 μm PES membrane stericup filters (Millipore Sigma) to remove cell debris and stored at −20°C until purification.

#### Protein purification

Protein purification by immobilized metal affinity chromatography (IMAC) and gravity flow was adapted from previous methods (23). After washing with Phosphate-Buffered Saline (PBS; Thermo Fisher Scientific), Nickel-Nitrilotriacetic acid (Ni-NTA) agarose (Qiagen) was added to culture supernatant followed by overnight incubation (12-16 hours) at 4 °C on a rotator. For every 150 mL of culture supernatant, 2.5 mL of Ni-NTA agarose was added. 5mL gravity flow polypropylene columns (Qiagen) were equilibrated with PBS. One polypropylene column was used for every 150 mL of culture supernatant. The supernatant-agarose mixture was then loaded onto the column to retain the agarose beads with recombinant proteins bound to the beads. Each column was then washed, first with 1X culture supernatant volume of PBS and then with 25 mL of 20 mM imidazole (Millipore Sigma) in PBS wash buffer to remove host cell proteins. Recombinant proteins were then eluted from each column in three fractions with 5 mL of 250 mM imidazole in PBS elution buffer per fraction giving a total of 15 mL eluate per column. The eluate was subsequently dialyzed several times against PBS using Amicon Ultra Centrifugal Filters (Millipore Sigma) at 7000 rpm for 20 minutes at 10 °C to remove the imidazole and concentrate the eluate. Filters with a 10 kDa molecular weight cut-off were used for S-RBD eluate. The final concentration of the recombinant S-RBD and S proteins was measured by bicinchoninic acid (BCA) assay (Thermo Fisher Scientific), and purity was assessed on 10% SDS-PAGE (Bio-Rad) followed by Coomassie blue staining. After sufficient destaining in water overnight, clear single bands were visible for S-RBD.

#### Viruses and cells

Vero-E6 cells (ATCC CRL-1586) and Vero-E6-TMPRSS2 cells (24) were cultured in Dulbecco’s modified Eagle medium (DMEMD) containing 10% fetal bovine serum (Gibco), 1 mM glutamine (Invitrogen), 1 mM sodium pyruvate (Invitrogen), 100 U/ml of penicillin (Invitrogen), and 100 μg/ml of streptomycin (Invitrogen) (complete media or CM). Cells were incubated in a 5% CO2 humidified incubator at 37°C. The SARS-CoV-2/USA-WA1/2020 virus was obtained from BEI Resources. The infectious virus titer was determined on Vero cells using a 50% tissue culture infectious dose (TCID50) assay as previously described for SARS-CoV (25, 26). Serial 10-fold dilutions of the virus stock were made in infection media (IM, which is identical to CM except the FBS is reduced to 2.5%), then then 100 μl of each dilution was added to Vero cells in a 96-well plate in sextuplicate. The cells were incubated at 37°C for 4 days, visualized by staining with naphthol blue-black, and scored visually for cytopathic effect. A Reed and Muench calculation was used to determine TCID50 per ml (27).

### Measurement of endpoint anti-S-RBD IgG titer

The protocol was adapted from a published protocol from Dr. Florian Krammer’s laboratory (Stadlbauer 2020). Ninety-six well plates (Immulon 4HBX, Thermo Fisher) were coated with S-RBD at a volume of 50 μl of 2 μg/ml of diluted antigen in filtered, sterile 1xPBS (Thermo Fisher) at 4°C overnight. Coating buffer was removed, plates were washed three times with 300 μl of PBS-T wash buffer (1xPBS plus 0.1% Tween 20, Fisher Scientific), and blocked with 200 μl of PBS-T with 3% non-fat milk (milk powder, American Bio) by volume for one hour at room temperature. All plasma samples were heat inactivated at 56°C on a heating block for one hour prior to use. Negative control samples were prepared at 1:10 dilutions in PBS-T in 1% non-fat milk and plated at a final concentration of 1:100. A monoclonal antibody (mAb) specific for the SARS-CoV-2 spike protein was used as a positive control (1:5,000, Sino Biological). For serial dilutions of plasma on S-RBD coated plates, plasma samples were prepared in three-fold serial dilutions starting at 1:20 in PBST in 1% non-fat-milk. Blocking solution was removed and 10 μl of diluted plasma was added in duplicates to plates and incubated at room temperature for two hours. Plates were washed three times with PBS-T wash buffer and 50 μl secondary antibody was added to plates and incubated at room temperature for one hour. Anti-human secondary antibodies used included Fc-specific total IgG HRP (1:5,000 dilution, Invitrogen), prepared in PBS-T plus 1% non-fat milk. Plates were washed and all residual liquid removed before adding 100 μl of SIGMAFAST OPD (o-phenylenediamine dihydrochloride) solution (Sigma Aldrich) to each well, followed by incubation in darkness at room temperature for ten minutes. To stop the reaction, 50 μl of 3M hydrochloric acid (HCl, Fisher Scientific) was added to each well. The OD of each plate was read at 490nm (OD490) on a SpectraMax i3 ELISA plate reader (BioTek). The positive cutoff value for each plate was calculated by summing the average of the negative values and three times the standard deviation of the negatives. All values at or above the cutoff value were considered positive. Values were graphed on a dose response curve, a best fit line drawn by nonlinear regression, and area under the curve (AUC) calculated.

### Measurement of endpoint neutralization titer

Plasma neutralization titers were determined as described for SARS-CoV (Schaecher 2008). Two-fold dilutions of plasma (starting at a 1:20 dilution) were made in IM. Infectious virus was added to the plasma dilutions at a final concentration of 1×104 TCID50/ml (100 TCID50 per 100ul). The samples were incubated for one hour at room temperature, then 100 uL of each dilution was added to one well of a 96 well plate of VeroE6-TMPRSS2 cells in sextuplet for 6 hours at 37°C. The inoculums were removed, fresh IM was added, and the plates were incubated at 37°C for 2 days. The cells were fixed by the addition of 150 uL of 4% formaldehyde per well, incubated for at least 4 hours at room temperature, then stained with Napthol blue black. The nAb titer was calculated as the highest serum dilution that eliminated cytopathic effect (CPE) in 50% of the wells. Values were graphed on a dose response curve, a best fit line drawn by nonlinear regression, and area under the curve (AUC) calculated.

### Cell staining and flow cytometry

PBMCs were isolated from blood using ficoll separation gradient and viably frozen. Cells were thawed before use, and 5e6-10e6 PBMCs were stained from each participant. Fc blocker (BD Cat #564220) diluted in FACS Buffer (1x PBS with 1% BSA) was added to the cells and incubated for 30minutes on ice or 4c. The cells were then washed twice with FACS buffer. Soluble 6X-His tagged S-RBD protein was then added to the cells and incubated at room temperature for 30 min. This was followed by wash steps with FACS buffer. Conjugated antibodies (Suppl. Table 1 for list of antibodies and their conjugate fluorophores) and live dead stain was then added to the cells and incubated for an additional 30min. The cells were washed two or three more times before running the cells on BD Biosciences LSR II instrument. 1e6-6e6 total events were collected for each participant, resulting in medians of 69.5 absolute RBD+ and 13,971 RBD-class switched MBC for each participant (range=1-454 RBD+, 152-84,645 RBD-MBC). For UMAP analysis, all RBD+ class switched MBC were included, and RBD-class switched MBC were downsampled to 3000 cells per subject. The subject with only 1 RBD+ cell was excluded from RBD+ MBC subset analyses. Positive gates for each fluorophore were set after compensation and using fluorescence minus one (FMO) staining and isotype control antibodies.

### Statistical analysis

FlowJo software was used to analyze all the flow results from the LSRII. Statistical analyses were performed in Prism (Graphpad software). Two group comparisons were performed with t tests if data were normally distributed based on Shapiro Wilk normality test or Mann Whitney rank test if data were not normally distributed. Multi-group comparisons were performed using one-way ANOVA, with p values adjusted for multiple comparisons using the Benjamini, Krieger and Yekutieli method.

### Study approval

This research was approved by the Johns Hopkins University School of Medicine’s Institutional Review Board (IRB). Prior to blood collection, all participants provided informed written consent.

## Data Availability

After publication, all data referred to in the manuscript is available on request.

## Author Contributions

COO conceived the project, performed experiments, interpreted data, and wrote the initial manuscript draft; NES conceived the project, performed experiments, and interpreted data; PWB interpreted data; HSP, KL, and AG performed experiments and interpreted data; AA interpreted data; SCR interpreted data; PL and MJB provided reagents. AP performed experiments and interpreted data; SLK performed experiments and interpreted data; YCM conceived the project and provided participant samples; ALC conceived the project, provided participant samples, and interpreted data; JRB conceived the project, interpreted data, and wrote the initial manuscript draft. All authors reviewed and edited the manuscript.

## Acknowledgements

We would like to thank participants in the study, Florian Krammer for providing the S-RBD plasmid, and members of the Johns Hopkins Viral Hepatitis Center for thoughtful discussion. We thank the Bloomberg Flow Cytometry and Immunology Core for equipment and technical assistance. We thank the National Institute of Infectious Diseases, Japan, for providing VeroE6TMPRSS2 cells and acknowledge the Centers for Disease Control and Prevention, BEI Resources, NIAID, NIH for SARS-related coronavirus 2, isolate USA-WA1/2020, NR-5228.This research was supported by the National Institutes of Health grants R01AI127469 and R21AI151353 (to J.R.B.), U54CA260492 (SLK and AC), and in part by the NIH Center of Excellence for Influenza Research and Surveillance (HHSN272201400007C, to AP and SLK)..

