## Supplemental Data for "Durable SARS-CoV-2 B cell immunity after mild or severe disease"

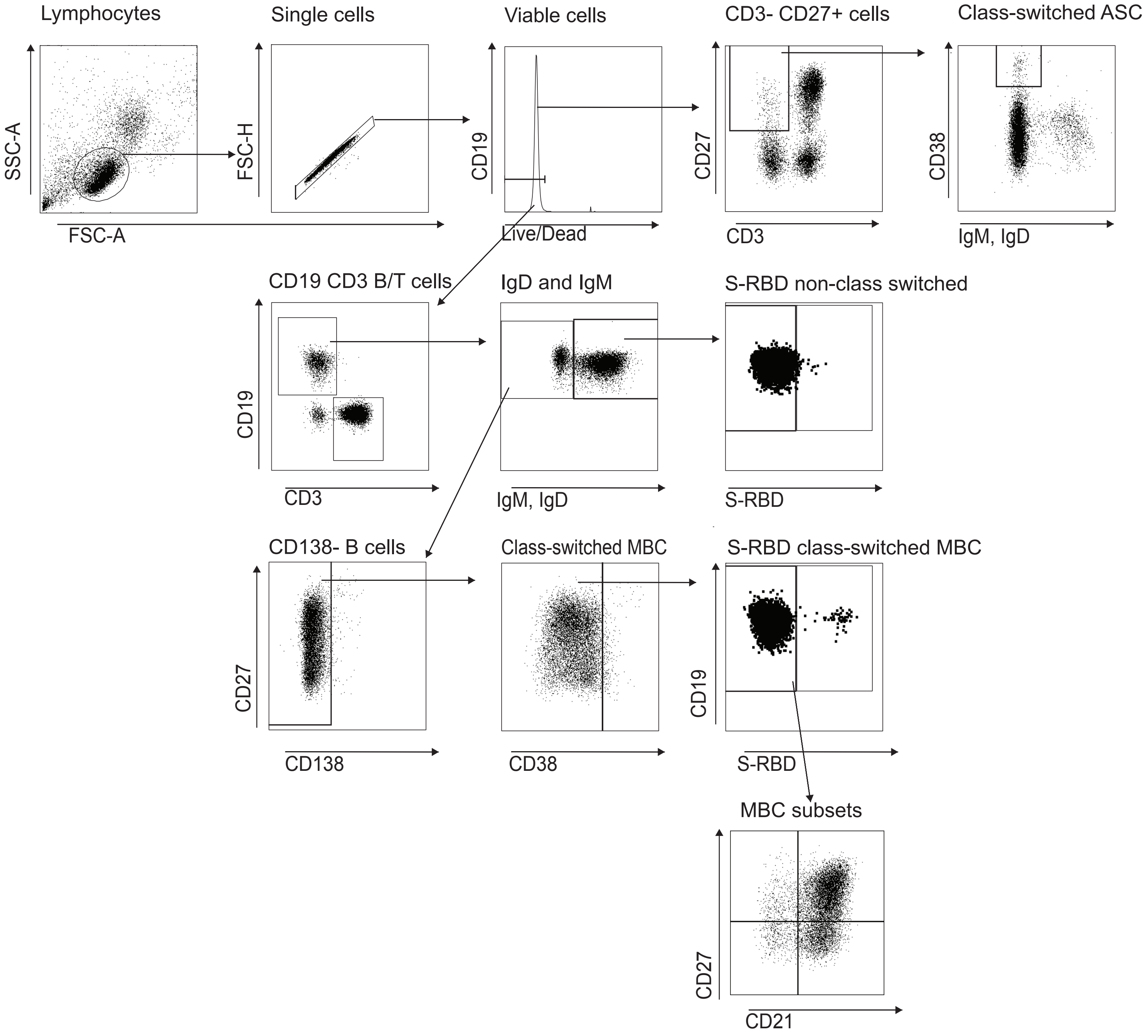


**Supplemental Figure 1: Gating schematic.** All antibodies used are listed in supplemental table 1. Fluorescence minus one (FMO) and isotype controls were used as guide to differentiate positive and negative populations. For S-RBD specific B cells, healthy participants were used as controls to set the gate for positive cells. All cells were run on an LSRII flow cytometer and analyzed using FlowJo.

**Supplemental Table 1. Antibodies used**

| Marker | Fluorophore | Clone | Cat # | Vendor |
|---|---|---|---|---|
| IgM | BB515 | G20-127 | 564622 | BD Biosciences |
| IgD | BB515 | IA6-2 | 565243 | BD Biosciences |
| CD19 | APC-R700 | HIB19 | 564977 | BD Biosciences |
| CD3 | APC-H7 | SK7 | 560176 | BD Biosciences |
| CD138 | BV711 | MI15 | 563184 | BD Biosciences |
| CD21 | PE-CF594 | B-LY4 | 563474 | BD Biosciences |
| CD27 | BV786 | L128 | 563327 | BD Biosciences |
| CD38 | BB700 | HIT2 | 566445 | BD Biosciences |
| BTLA (CD272) | BV421 | J168-540 | 564802 | BD Biosciences |
| CD22 | PE-Cy7 | HIB22 | 563941 | BD Biosciences |
| FCRL5 (CD307e) | PE | 509F6 | 566734 | BD Biosciences |
| CXCR5 (CD185) | BV480 | RF8B2 | 566142 | BD Biosciences |
| 6x-His Tag mAb | Alexa Fluor 647 | HIS.H8 | MA1-21315-A647 | ThermoFisher |
| Fixable Yellow Dead | BV480 | Viablity Dye | L34968 | ThermoFisher |
| IgG1 Kpa ItCl | BB700 | X40 | 566404 | BD Biosciences |
| IgG1 Kpa ItCl | BV711 | X40 | 563044 | BD Biosciences |
| IgG1 Kpa ItCl | BV421 | X40 | 562438 | BD Biosciences |
| IgG1 Kpa ItCl | PE-Cy7 | MOPC-21 | 557872 | BD Biosciences |
| IgG2a Kpa ItCl | PE | G155-178 | 554648 | BD Biosciences |
| Rat/Ham Ig Kpa Comp Bead | | | 552845 | BD Biosciences |
| Ms Ig Kpa Comp Bead Set | | | 552843 | BD Biosciences |
| Hu Fc Block Pure Fc1.3216 | | | 564220 | BD Biosciences |
| ArC Amine Comp | | | A10346 | ThermoFisher |
